# Post-stroke depression, but not anxiety, is independently associated with executive and visuospatial function: a five-year longitudinal cohort study

**DOI:** 10.64898/2026.08.04.26359690

**Authors:** Florine Ruthmann, Etienne Allart, Anne-Marie Mendyk Bordet, Dominique Deplanque, Régis Bordet, Thibaut Dondaine

**Author notes:** **Corresponding author:** Thibaut Dondaine, PhD Service de pharmacologie, Faculté de Médecine, Pôle Recherche, 1, place de Verdun 59045 Lille CEDEX.

## Abstract

**Background:** Post-stroke anxiety and depression frequently co-occur, and whether each is independently associated with cognitive impairment remains unclear because most studies model one without adjusting for the other variable. In a five-year cohort, we tested whether anxiety and depression have distinct cognitive correlates and whether early affective status predicted subsequent cognitive recovery.

**Methods:** Patients from the STROKDEM cohort were assessed at 6, 12, 36, and 60 months post-stroke for anxiety, depression, and five cognitive domains (memory, executive functioning, attention, visuospatial functioning, and language). Two symmetric random-intercept linear mixed models regressed each affective score on the cognitive domains while adjusting for other scores. Repeated-measures correlations, multiple imputations for attrition, and exploratory trajectory and prognostic models were also used.

**Results:** After mutual adjustment and correction, depression was independently associated with executive and visuospatial function, whereas anxiety showed no independent cognitive correlation. Repeated-measures correlation confirmed this dissociation. The anxiety findings were stable across the sensitivity analyses and multiple imputations. Attrition was selective for baseline cognition, and exploratory associations between early affect and cognitive recovery did not survive multiple imputations and were inconclusive.

**Limitations:** Attrition was substantial and selective on baseline cognition; the persistent anxiety subgroup was small, limiting the power for trajectory analyses; and psychiatric history, psychotropic medication, and cognitive reserve beyond education were unavailable.

**Conclusions:** The cognitive burden of post-stroke affective disorders is carried by depression, rather than anxiety. Because anxiety-related cognitive impairment largely reflects comorbid depression, screening for depression rather than anxiety alone may better identify stroke survivors at risk of cognitive impairment.

## Introduction

There is growing interest in the cognitive and psychiatric consequences of stroke; however, research has overwhelmingly focused on post-stroke depression (PSD) (Robinson and Jorge, 2016), while post-stroke anxiety (PSA) remains underdiagnosed and understudied. One-fifth to one-quarter of stroke survivors experience clinically significant anxiety (Knapp et al., 2020), which affects rehabilitation participation, recovery, and quality of life (Donnellan et al., 2006). Because PSA has often been overlooked, comparatively little is known about its trajectory, determinants, and cognitive correlates.

PSA is most often studied in the subacute phase, where it largely reflects the reaction to the traumatic event of stroke. However, it can become chronic, with a high prevalence reported years later (Åström, 1996; Bergersen et al., 2010), and early- and late-onset PSA may not share the same determinants (Castillo et al., 1995; Lee et al., 2019; Lincoln et al., 2013). There is now a recognized need to describe the longitudinal course of PSA over an extended follow-up and to distinguish patients with persistent anxiety from those with inconsistent anxiety, rather than relying on a single assessment (Oosterveer et al., 2026).

A central difficulty in interpreting the cognitive correlates of PSA is its comorbidity with depression. PSA and PSD are highly prevalent and strongly correlated (Menlove et al., 2015), and each has been independently linked to fatigue and sleep disturbance (Galligan et al., 2016; Sanner Beauchamp et al., 2020; Wright et al., 2017). Cognitive disorders are common after stroke (Sun et al., 2014), and anxiety disorders have well-established cognitive components. In both general and clinical populations, anxiety has been associated with executive and attentional impairment (Airaksinen et al., 2005; Castaneda et al., 2008; Hallion et al., 2017), even in tasks unrelated to threat (Balderston et al., 2017). Therefore, one might expect PSA to be accompanied by a distinctive cognitive profile after stroke.

However, the evidence is inconsistent, and the inconsistency largely depends on whether depression is considered. PSA is associated with cognitive complaints (van Rijsbergen et al., 2020), but whether it is associated with poorer objective performance remains debated (Barker-Collo, 2007; Castillo et al., 1995; Lo Buono et al., 2018). Williams and Demeyere (2021) reported that anxiety was associated with global cognition and visuospatial ability, but the association disappeared when depression was controlled for. In contrast, a work by our team with a pooled analysis of three cohorts from the STROKOG consortium found anxiety to be associated with global cognitive impairment even after adjustment for depression (Ruthmann et al., 2025). A further longitudinal study found that long-term anxiety was related to subjective complaints but not to objective global performance (Kusec et al., 2024). These divergent results share a methodological root: anxiety and depression are rarely modelled symmetrically, so an apparent cognitive correlate of one may, in fact, belong to the other.

Resolving this question requires three features rarely combined in a single study: repeated assessment over a long follow-up period, a domain-structured neuropsychological battery rather than a global score, and a design that adjusts each affective dimension for the other so that their cognitive correlates can be distinguished rather than confounded.

The present study used a five-year STROKDEM cohort to address this gap. Our primary aim was to determine whether PSA and PSD have distinct domain-specific cognitive correlates when modelled symmetrically. A secondary, exploratory aim was to test whether early affective status predicts subsequent cognitive recovery. Given the comorbidity of the two conditions, we examined whether the cognitive correlates commonly attributed to anxiety would, once depression is accounted for, be carried primarily by depression.

## Methods

### Participants and design

This work is ancillary to the STROKDEM study (NCT01330160), conducted only at Lille Hospital, which aimed to investigate the prevalence and predictors of post-stroke dementia. The study was conducted between 2011 and 2018 and was approved by the local institutional review board (Comité de Protection des Personnes Nord Ouest IV, Lille, France; reference: 2009-A00141-56, March 17, 2009). Some data from this cohort have been the subject of several previous studies (Betrouni et al., 2022; Ruthmann et al., 2025).

The inclusion criteria were patients aged >18 years, those who provided informed consent, and those who had experienced a stroke less than 72 h prior. The exclusion criteria were as follows: pregnancy, malformations or traumatic cerebral hemorrhage, pure meningeal or intraventricular hemorrhage, guardianship or curatorship, contraindications to MRI or MRA, patients with no fluency in French or with no informant (resulting in inability to undergo neuropsychological testing), and pre-existing dementia. Pre-existing cognitive status was assessed using the Informative Questionnaire on Cognitive Decline in the Elderly (IQ-CODE). Patients with major cognitive impairment before stroke (IQ-CODE > 104) were excluded. The patients’ medical histories and baseline treatments are available in Supplementary Data (Table S1).

At inclusion, patients’ medical history and cardiovascular risk factors were collected. Patients were examined at 6, 12, 36, and 60 months after stroke for a comprehensive clinical examination, including neurological, functional, psychiatric, and cognitive evaluation.

This study included 202 patients. Eleven patients with hemorrhagic stroke were excluded from the analyses, resulting in 191 patients. Patients were lost during follow-up, and 85 patients were evaluated 5 years after stroke.

### Measures

A neurologist confirmed stroke using computed tomography (CT), magnetic resonance imaging (MRI), or other methods. Data on stroke type, stroke side, biological sex, age, education level, and medical history were collected as well. Ischemic stroke types were categorized according to the TOAST classification (Adams et al., 1993). Stroke severity was determined using the NIHSS. The Index for Activities of Daily Living (IADL) was used to measure the dependency.

### Neuropsychological evaluation

#### Cognitive Evaluation

A comprehensive cognitive assessment was conducted at each time point. Global cognition was assessed using the MoCA and MMSE. Memory, executive function, attention, language, and visuospatial abilities were also evaluated (Table 1). Cognitive scores were expressed as the average Z-scores per domain, as in previous studies (Lo et al., 2023).

**Table 1.**
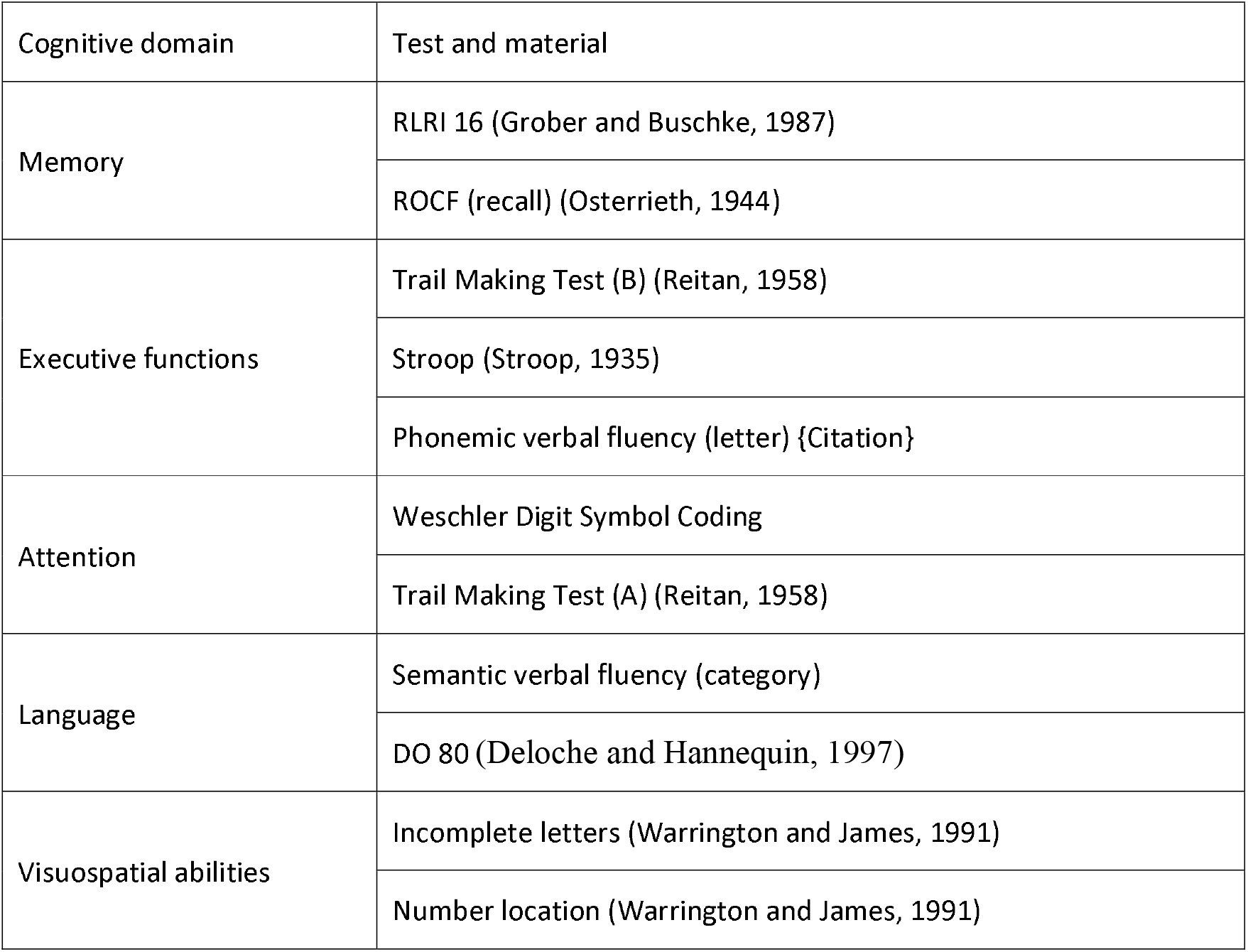
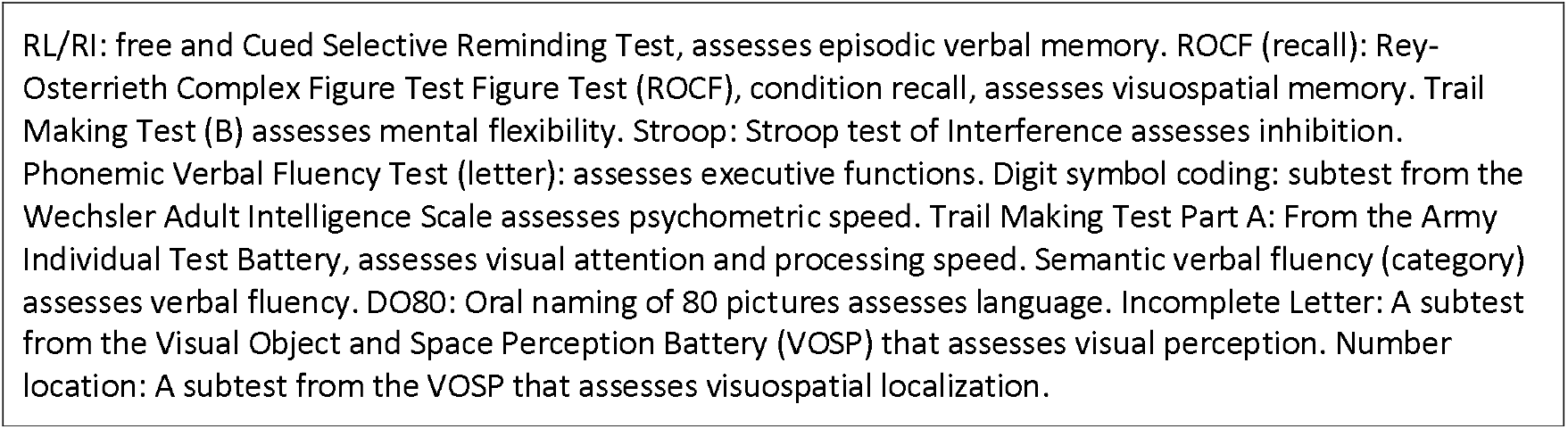
Tests employed for each cognitive domain.

#### Psychiatric evaluation

The Hamilton Anxiety Rating Scale (HARS) provides an anxiety score with a cut-off of ≥ 8 to detect a significant level of anxiety, as recommended by Matza et al. (Matza et al., 2010). The Center for Epidemiologic Studies Depression Scale (CES-D) provides a depression score, with a cut-off of ≥16 recommended to detect a significant level of depression (Eaton, 2004). The Chalder Fatigue Scale provides a score for physical and psychological fatigue, with a score ≥ 4 recommended to indicate significant fatigue (Chalder et al., 1993).

### Statistical analysis

Analyses were conducted in R 4.5.3, with a pre-specified hierarchy of primary, secondary, and exploratory analyses. Because affective and cognitive measures were repeated within patients, all longitudinal models included a random intercept for patients to account for the non-independence of repeated observations. HARS and CES-D scores were right-skewed with a floor at zero and were square-root transformed, which brought model residuals close to normality; residuals were inspected on quantile–quantile (Q-Q) plots. No observation was excluded on the basis of its residuals, so that the full range of affective symptoms was retained.

The primary analysis comprised two symmetric random-intercept linear mixed models. First, square-root-transformed anxiety was regressed on time, sex, age, education, the five cognitive domain z-scores, stroke severity (NIHSS), and depression. Second, depression was regressed on the same predictors, with anxiety substituted for depression. Modelling each affective dimension while adjusting for the other allowed their cognitive correlates to be distinguished rather than confounded by their comorbidity. Fixed effects were tested using Type II F tests with Satterthwaite denominator degrees of freedom, and multicollinearity was assessed using variance inflation factors (VIF). Within each model, the p-values for the five cognitive domains were adjusted for the false discovery rate (FDR) using the Benjamini– Hochberg procedure.

The robustness of the primary models was examined through pre-specified sensitivity analyses: refitting on untransformed and log-transformed scores; a bounded-influence robust mixed model; a subject-level cluster bootstrap (2,000 resamples), which assumes neither normality nor homoscedasticity of residuals; refitting after excluding observations exceeding Cook’s distance of 4/n; and restriction to patients still assessed at 60 months. Because attrition was related to baseline cognition, missingness was further addressed by multiple imputation under a missing-at-random assumption (50 imputations; two-level predictive mean matching with patients as clustering variables; estimates pooled by Rubin’s rules) and by a pattern-mixture sensitivity analysis in which imputed post-dropout anxiety scores were shifted across a range of departures from missing-at-random.

As secondary analyses, within-patient associations between each affective score and cognition were estimated using repeated-measures correlations, reported both unadjusted and adjusted for the other affective scores, with Benjamin–Hochberg correction within each set of tests.

The patients were divided into three groups according to their anxiety scores. Patients were categorized as having persistent, fluctuating, or no anxiety. Persistent anxiety was defined as significantly high anxiety scores during the follow-up period. Fluctuating anxiety corresponded to patients who showed both non-anxious and anxious profiles during follow-up. Finally, non-anxious patients did not exhibit significant anxiety scores at follow-up.

Two sets of exploratory analyses were conducted in this study. First, patients with at least two available anxiety assessments were classified as never anxious, fluctuating, or persistently anxious, and the longitudinal course of each cognitive domain was modelled as a function of time, trajectory group, and their interaction within a mixed model, rather than a repeated-measures ANOVA, so that all available observations contributed, given the small persistent anxiety subgroup. Second, we tested whether early (M6) anxiety and depression predicted subsequent cognitive recovery (M12–M60): for each domain, the follow-up z-score was modelled as a function of time in interaction with the baseline affective score, adjusting for the baseline cognitive score in that domain, the other baseline affective score, age, sex, and education. These prognostic models were fitted to the observed data and re-estimated using multiple imputations. Two-sided p-values < 0.05 were considered significant.

## Results

### Sample and attrition

The number of available anxiety assessments declined from 157 at M6 to 139, 122, and 85 at M12, M36, and M60. Patients assessed at 60 months differed at baseline from those subsequently lost to follow-up: they had higher education (12.1 vs. 10.7 years, p = 0.021), lower functional disability (Rankin 0.73 vs. 1.10, p = 0.016), and better cognitive performance (MMSE 28.3 vs. 27.2, p = 0.003; MoCA 26.5 vs. 25.4, p = 0.044; executive z-score −0.43 vs. −0.82, p = 0.029; attention −0.33 vs. −0.90, p = 0.021). Baseline anxiety and depression did not differ between the two groups (both p > 0.22). Attrition was therefore selective on baseline cognition, which motivated the multiple imputation and pattern-mixture analyses reported below. The number of patients with available data at each time point is shown in Figure 1.

**Figure 1.**
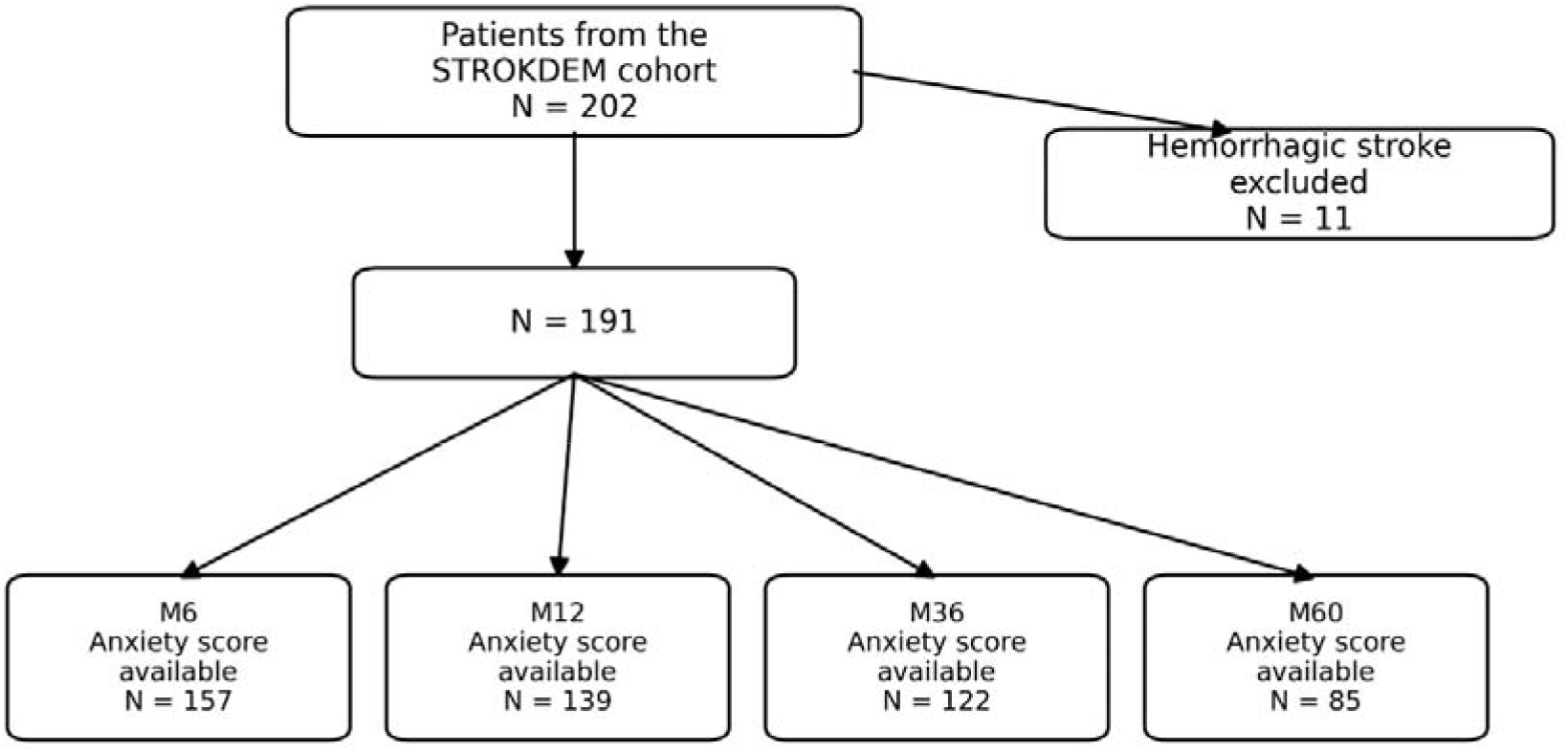
Number of patients with available anxiety score at each timepoint. **Note**. The cohort included 202 patients that were admitted to Lille Hospital for stroke. We excluded 11 patients with haemorrhagic stroke, resulting in 191 patients. Among them, 157 had available neuropsychological evaluation at the six-months visit, 139 at the twelve-months evaluation, 122 at the thirty-six-months evaluation and 85 at the sixty-months evaluation.

### Prevalence over five years

Anxiety persisted across the follow-up period, with a prevalence of 28.7% at M6, 26.6% at M12, 36.1% at M36, and 31.8% at M60. Depression followed the opposite course, declining from 27.1% at M6 to 29.5%, 22.1%, and 17.8%. Consistent with these trajectories, time was a strong predictor of depression in the mixed models (F(3,332) = 18.9, p < 0.001), but only weakly associated with anxiety (F(3,344) = 2.9, p = 0.037). All results are presented in Table 2.

**Table 2.** Prevalence of anxiety and depression and affective, clinical, and neuropsychological measures at each post-stroke assessment.

|  | **M6** | **M12** | **M36** | **M60** |
| --- | --- | --- | --- | --- |
| N assessed | 157 | 139 | 122 | 85 |
| ***Prevalence*** |  |  |  |  |
| Anxiety, n/N (%) [HARS $\geq 8$] | 45/157 (28.7) | 37/139 (26.6) | 44/122 (36.1) | 27/85 (31.8) |
| Depression, n/N (%) [CES-D $\geq 16$] | 42/155 (27.1) | 41/139 (29.5) | 27/122 (22.1) | 13/73 (17.8) |
| ***Affective and motivational*** |  |  |  |  |
| HARS (anxiety) | 6.34 (4.53) | 5.63 (5.26) | 6.75 (5.73) | 6.38 (7.21) |
| CES-D (depression) | 11.74 (8.26) | 11.29 (9.70) | 9.58 (9.87) | 7.79 (8.52) |
| LARS (apathy) | -27.51 (6.14) | -27.33 (6.71) | -28.34 (6.52) | -27.67 (7.84) |
| CFS (fatigue) | 3.98 (3.20) | 3.63 (3.15) | 3.44 (3.23) | 3.27 (2.93) |
| ***Clinical*** |  |  |  |  |
| NIHSS | 0.77 (1.60) | 0.63 (1.65) | 0.56 (1.34) | 0.50 (1.21) |
| mRS | 0.92 (1.03) | 0.85 (1.01) | 0.88 (1.01) | 1.02 (1.11) |
| IADL | 12.37 (2.70) | 12.49 (2.39) | 12.73 (1.88) | 12.63 (2.13) |
| ***Global cognition*** |  |  |  |  |
| MMSE (/30) | 27.75 (2.50) | 27.88 (2.68) | 27.86 (2.76) | 27.70 (3.25) |
| MoCA (/30) | 25.97 (3.52) | 26.41 (3.74) | 26.69 (3.19) | 26.64 (3.23) |
| ***Cognitive domains (z-scores)*** |  |  |  |  |
| Memory | -0.13 (0.95) | -0.07 (0.93) | -0.03 (0.95) | -0.20 (1.00) |
| Executive | -0.61 (1.10) | -0.74 (1.35) | -0.45 (1.39) | -0.06 (0.85) |
| Attention | -0.60 (1.49) | -0.42 (1.19) | -0.29 (1.33) | -0.14 (1.12) |
| Visuospatial | 0.03 (0.77) | 0.06 (0.70) | 0.18 (0.77) | 0.39 (0.96) |
| Language | -0.28 (0.91) | -0.12 (0.84) | -0.07 (0.75) | -0.10 (0.72) |

Values are expressed as mean (SD), unless otherwise indicated. Anxiety was defined as HARS $\geq 8$ and depression as CES-D $\geq 16$ . HARS, Hamilton Anxiety Rating Scale; CES-D, Center for Epidemiologic Studies Depression Scale; LARS, Lille Apathy Rating Scale; CFS, Chalder Fatigue Scale; NIHSS, National Institutes of Health Stroke Scale; mRS, modified Rankin Scale; IADL, Instrumental Activities of Daily Living; MMSE, Mini-Mental State Examination; MoCA, Montreal Cognitive Assessment. Cognitive domains are expressed as standardized z-scores. The apparent improvement in cognitive z-scores
over time partly reflects selective attrition, as patients assessed at later time points had better baseline cognition (see Results).

### Cross-sectional cognitive correlates: the anxiety–depression contrast

At M6, anxious patients had higher depression (CES-D 17.1 vs. 9.6, p < 0.001), more fatigue (CFS 5.4 vs. 3.3, p < 0.001), poorer functional autonomy (IADL 11.7 vs. 12.9, p = 0.031), and lower scores in several cognitive domains in unadjusted comparisons (memory, executive, language; all p < 0.03). However, these unadjusted differences do not separate anxiety from comorbid depression.

The two symmetric mixed models separated them, and the contrast was clear-cut. After mutual adjustment and FDR correction, anxiety had no independent cognitive correlate; no domain reached significance (memory p_FDR = 0.80; executive 0.64; attention 0.34; visuospatial 0.80; language 0.079), with language association being the only near-significant effect (b = −0.185, 95% CI [−0.33, −0.04]). Depression, in contrast, was independently associated with two domains: executive function (b = −0.167, 95% CI [−0.28, −0.05], p_FDR = 0.021) and visuospatial abilities (b = −0.208, 95% CI [−0.38, −0.04], p_FDR = 0.043). In each model, the other affective score was by far the strongest predictor (depression in the anxiety model, F(1,384) = 145.8; anxiety in the depression model, F(1,451) = 127.8; both p < 0.001). Multicollinearity was negligible (all VIF < 1.5), and the intraclass correlations (0.34 and 0.52) supported the mixed model specification.

The repeated-measures correlations (r_rm) reproduced this dissociation using an independent method. After adjusting for anxiety, depression remained associated with executive functioning (r_rm = −0.19, p_FDR = 0.004) and visuospatial abilities (r_rm = −0.13, p_FDR = 0.038). After adjusting for depression, anxiety was not associated with any cognitive domain (closest: attention, p_FDR = 0.12). Both affective scores were strongly and independently associated with fatigue and apathy (all p_FDR ≤ 0.02).

Sex was not associated with anxiety or depression levels in either mixed model (both p > 0.19), nor did it modulate cognitive correlates. In exploratory analyses, women were more likely than men to experience at least one anxious episode over the follow-up (63.5% vs. 45.2%), and the point prevalence of anxiety was higher in women (36.3% vs. 27.4%), but both differences were borderline (p = 0.052), consistent with the limited power afforded by the cohort’s male-predominant sex ratio (119 men, 72 women).

**Figure 2.**
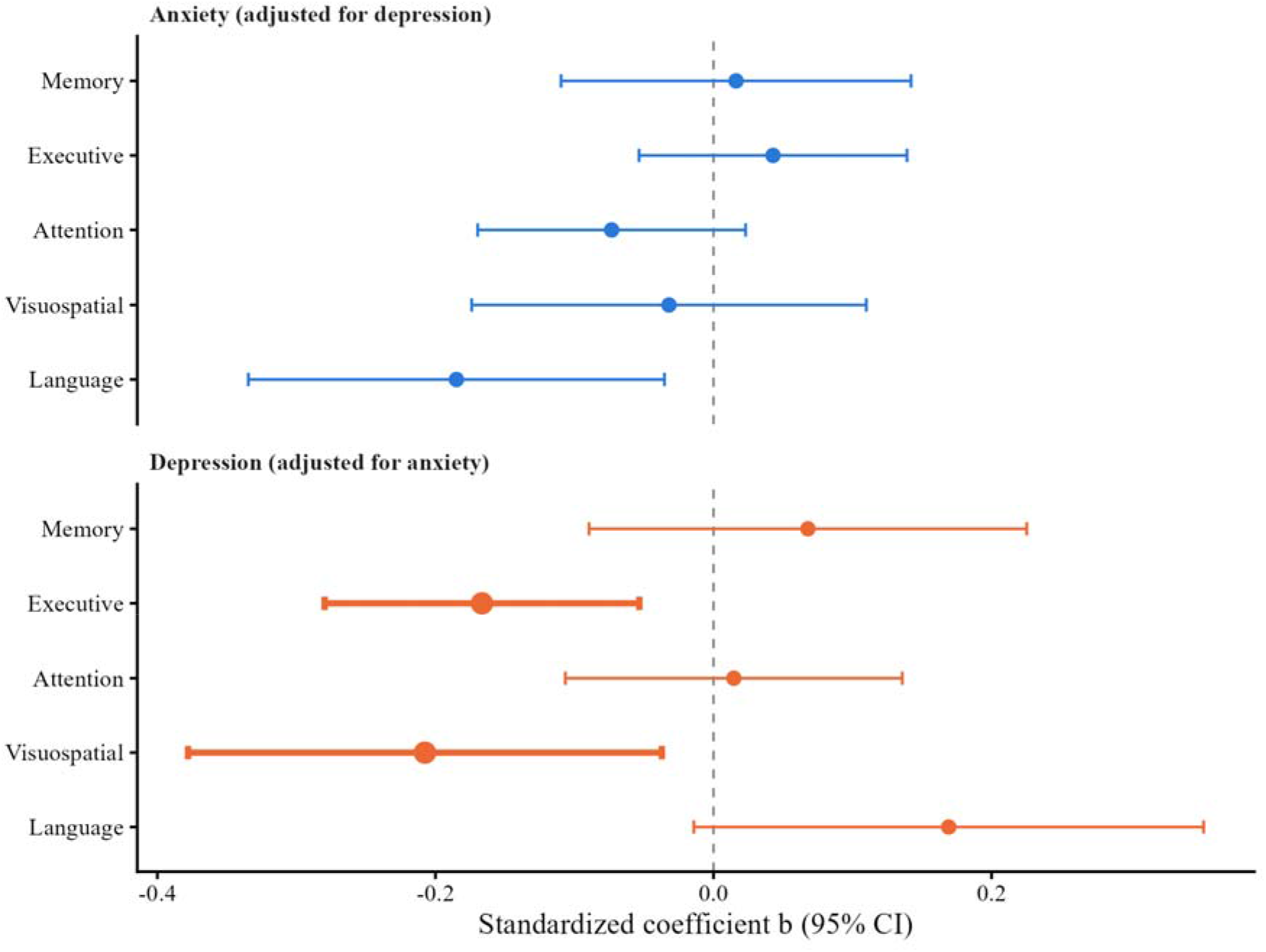
Cognitive correlate of anxiety and depression, mutually adjusted. Standardized coefficients (b) with 95% confidence intervals from two symmetric random-intercept mixed models, each affective score adjusted for the other and for time, sex, age, education and stroke severity. Thicker intervals denote domains significant after Benjamini–Hochberg correction: executive (depression, p_FDR = 0.021) and visuospatial (depression, p_FDR = 0.043).

### Robustness of the anxiety findings

The absence of an independent anxiety–cognition association was stable across all sensitivity analyses. The borderline language effect was significant only in the specifications most sensitive to the distribution’s tail (untransformed p = 0.073, log p = 0.011, robust p = 0.018, exclusion of influential observations p = 0.047) and in the cluster bootstrap (b = −0.187, 95% CI [−0.342, −0.047]); however, it did not survive restriction to completers (p = 0.098) and, critically, was abolished under multiple imputation (b = −0.102, p = 0.106). The pattern-mixture analysis was flat: across shifts of −0.5 to +0.5 SD in post-dropout anxiety, no cognitive domain was significant. The depression covariate remained highly significant throughout the study (imputed b = 0.054, p < 0.001).

### Anxiety trajectories (exploratory)

Patients were classified as never anxious (n = 70), fluctuating (n = 59), or persistently anxious (n = 16). Cognitive performance improved over time across domains (time effect significant for executive, attention, visuospatial, and language, all p_FDR < 0.03), but neither the group main effect nor the time-by-group interaction survived correction (smallest interaction p_FDR = 0.097, executive). Available data were sparse in the persistent group (16, 14, 9, and 8 patients across the four time points; only seven with all four assessments), so these trajectory analyses were underpowered and reported as hypothesis-generating.

### Early affect and cognitive recovery (exploratory)

We tested whether early affective status predicted the rate of cognitive recovery over the M12–M60 period. In the observed data, both early anxiety (executive recovery: time-by-anxiety interaction F(2,175) = 5.19, p_FDR = 0.032) and early depression (visuospatial: F(2,202) = 7.30, p_FDR = 0.004; language: F(2,194) = 5.08, p_FDR = 0.018) appeared to predict recovery slopes. However, these effects did not survive multiple imputations: under pooling, none of the interaction terms remained significant (early anxiety on executive recovery, longest-interval term p = 0.68; early depression on visuospatial and language recovery, longest-interval terms p = 0.23 and p = 0.22). Given that attrition was selective on baseline cognition and that only approximately 36 anxious and 35 depressed patients had any follow-up cognitive assessment, these complete-case associations most plausibly reflect the missing-data mechanism rather than a genuine prognostic effect. Therefore, we treated the prognostic analyses as exploratory and inconclusive.

## Discussion

In a five-year cohort with domain-specific neuropsychological assessment, we asked whether post-stroke anxiety and depression have distinct cognitive correlates after adjusting for the other. The answer is a clear dissociation, but not the one most often assumed: when anxiety and depression were modelled symmetrically, the cognitive signal was carried by depression, which was independently associated with executive and visuospatial performance, whereas anxiety had no cognitive correlate. Two independent analytic approaches (symmetric mixed models and partial repeated-measures correlations) converged on this conclusion, which was robust to transformation, influential observations, and handling of missing data.

This result reconciles the apparently contradictory literature. Williams and Demeyere (2021) found that anxiety was associated with global and spatial attention cognition at six months, but these associations did not remain significant after adjustment for co-occurring depression, whereas depression was related to every cognitive domain assessed. Our findings extend theirs to a five-year horizon and a domain-structured longitudinal design and reach the same conclusion. Convergent evidence has been rapidly accumulated. Using the same Oxford Cognitive Screen, Milosevich et al. (2025) found domain-specific impairment associated with depression but not anxiety at six months. Kelleher et al. (2026), after adjusting for anxiety and education and corroborating their findings in multiply imputed data, reported that within-domain impairments (including visuospatial attention, episodic memory, and picture naming) uniquely predicted depression severity. The domains they identified closely mirror the executive and visuospatial correlates observed in the present study. The pooled STROKOG analysis, which retained an anxiety–cognition association after adjustment for depression (Ruthmann et al., 2025), differs in using a single global cognitive outcome rather than domain scores. The divergence between studies is best explained by whether depression is modelled symmetrically and whether cognition is treated globally or by domains. Our data suggest that much of the cognitive impairment previously attributed to anxiety reflects its comorbidity with depression.

Dissociation is also interpretable at the domain level. The two domains independently linked to depression (executive functioning and visuospatial ability) are among those most consistently implicated in post-stroke depression and in the vascular depression framework, in which frontosubcortical and right-hemisphere networks are affected. Earlier work has already pointed to executive functioning and working memory as the strongest cognitive correlates of post-stroke depression (Hommel et al., 2015), and recent large-scale data reinforce the specificity: in over a thousand patients, depression independently predicted impairment across executive, visuospatial, and language domains after adjustment (Cankaya et al., 2025). The absence of an independent anxiety correlate, even in the attentional domain where anxiety-related biases are classically expected (Eysenck et al., 2007), indicates that in a stroke population, the shared variance of the two affective conditions, rather than anxiety-specific mechanisms, drives the observed cognitive associations.

### Prevalence and trajectory

Anxiety was common and persistent, affecting 28.7% of patients at six months and remaining between one-quarter and one-third throughout the five-year follow-up, whereas depression declined over the same period. Persistent anxiety during the chronic phase is consistent with previous long-term reports (Bergersen et al., 2010; Lincoln et al., 2013). This contrast between conditions has been reported before: in a two-year follow-up, recovery from anxiety and depression was high (around 80%), whereas apathy remained stable (Sagen-Vik et al., 2022), underscoring that affective sequelae follow distinct courses. When patients were classified by longitudinal profile, roughly half showed fluctuating anxiety and a minority showed persistent anxiety (Oosterveer et al., 2026), supporting the robustness of these trajectory groups. The contrast between a stable anxiety prevalence and a declining depression prevalence is clinically relevant, as it implies that anxiety, more than depression, is the affective symptom that endures and may therefore be under-addressed in long-term care.

### Clinical and psychological correlates

Anxiety was not associated with stroke severity or functional independence, in line with the view that post-stroke psychiatric symptoms do not simply track lesion burden (Ayerbe et al., 2011). Our cohort had a relatively mild clinical profile (Ruthmann et al., 2025), which should be considered. Both anxiety and depression were strongly correlated with fatigue and apathy, but neither fatigue (entered as a covariate) nor apathy (omitted to avoid collinearity with depression) accounted for cognitive findings. The strong and expected anxiety–depression comorbidity (Menlove et al., 2015) is precisely what motivated the symmetric modelling strategy and what makes the resulting dissociation informative rather than an artifact of confounding. The higher rate of anxious episodes in women, although reaching only borderline significance here, is consistent with converging recent literature: independently of stroke severity, age, and education, women show higher anxiety across the first post-stroke year (Duttagupta et al., 2025), and a markedly higher point prevalence of anxiety has been reported in female survivors (Suñer-Soler et al., 2024).

### Prognostic analyses and the role of attrition

We hypothesized that early affective status might shape subsequent cognitive recovery as a plausible expectation, since early cognition and mood have each been shown to predict later affective and cognitive outcomes (Dec-Ćwiek et al., 2025), and baseline vascular cognitive impairment predicts the later course of apathy, though notably not of depression (Douven et al., 2016). In our complete-case analyses, early depression appeared to predict poorer visuospatial and language recovery, and early anxiety predicted poorer executive recovery.

However, none of these interactions survived multiple imputations, and attrition in this cohort was demonstrably selective for baseline cognition. The most parsimonious interpretation is that complete-case prognostic associations were generated by informative dropouts rather than by a genuine effect of early affect on recovery. Therefore, we report these analyses as exploratory and inconclusive. This caution is not merely local: a ten-year trajectory study of post-stroke depression explicitly acknowledged that its findings were confined to patients with repeated assessments, who had less severe disability, and might not generalize (Liu et al., 2023). In long-term stroke cohorts with cognition-related attrition, prognostic interactions estimated on completers alone may be seriously misleading, and multiple imputations should be regarded as a minimum safeguard rather than a refinement.

### Clinical implications

The practical message is one of screening. Because the cognitive burden associated with post-stroke affective disorders is carried by depression rather than anxiety, systematic screening for depression rather than anxiety alone is the more informative route for identifying stroke survivors at cognitive risk. Moreover, depression is the affective disorder most consistently linked to poorer long-term functional outcomes across domains of dependence, daily living, and cognition, from one month to five years post-stroke (Butsing et al., 2024), which strengthens the case for prioritizing its detection. This does not diminish the importance of anxiety, which is prevalent, persistent, and detrimental to quality of life and rehabilitation participation in its own right; rather, it clarifies that the cognitive dimension of long-term affective morbidity should be anchored in depression. Regular, domain-specific neuropsychological assessment alongside mood screening would allow rehabilitation and pharmacological strategies to be adjusted to the individual’s profile.

### Strengths and limitations

The main strengths of this study are the five-year follow-up, domain-structured cognitive battery, and an analytic strategy that models anxiety and depression symmetrically, corrects for multiplicity, and addresses attrition through multiple imputation and pattern-mixture sensitivity analyses. However, several limitations temper these findings. We did not have access to patients’ psychiatric histories and could not distinguish between pre-existing and post-stroke anxiety. We could not adjust for psychotropic medication, although most patients did not receive effective anxiolytic treatment. Cognitive reserve was measured only by education. Attrition was substantial and, at M60, was compounded by the COVID-19 pandemic. Although mixed models and multiple imputation use all available data under a missing-at-random assumption, unmeasured departures from that assumption cannot be excluded, which is precisely why prognostic analyses are treated as inconclusive. The effect sizes for the depression correlates were modest, and the persistent anxiety subgroup was small, limiting the power of the trajectory analyses. The cohort was male-predominant (119 men, 72 women), which is not representative of the broader stroke population and limits both the power to detect sex differences (the higher rate of anxious episodes in women reached only borderline significance) and the generalizability of sex-related findings. Finally, the methodology and materials remain a challenge in all stroke cohort studies because of the lack of testing that is designed specifically for stroke patients.

### Conclusion

Over five years post-stroke, the cognitive correlates of affective morbidity were carried by depression, not anxiety: depression was independently associated with executive and visuospatial function, whereas anxiety had no independent cognitive correlate once depression was considered. Distinguishing between the two conditions requires joint modelling; failing to do so risks misattributing depression-related cognitive impairment to anxiety. For clinical practice, screening for depression is the more informative route for identifying stroke survivors at cognitive risk, while prevalent and persistent anxiety warrants attention in its own right.

## Data Availability

All data produced in the present study are available upon reasonable request to the authors

## AUTHORS CONTRIBUTION

- Conceptualization: RB, TD, DD
- Data curation: A-MB
- Formal analysis: FR, TD
- Funding acquisition: RB
- Investigation: RB, TD, A-MB, EA, DD
- Methodology: TD, FR
- Writing – original draft: FR
- Writing – review and editing: all authors

## DATA AVAILABILITY

Data may be shared with researchers upon reasonable request, depending on the specific constraints of each cohort and research center.

## FUNDINGS

This study was funded by the Hauts-de-France Regional Council.

## Disclosure

The authors report no relevant disclosures.

---

**Table S1.** Patients’ medication 6 months after a stroke.

|  | Anxious patients | Non-anxious patients | Patients not evaluated for | Total |
| --- | --- | --- | --- | --- |
| Beta blocker | 16 | 36 | 6 | 58 |
| Anxiolytics | 13 | 7 | 3 | 23 |
| Hypnotics | 7 | 9 | 3 | 19 |
| Anti-depressant | 14 | 11 | 5 | 30 |
| Antiepileptic | 5 | 6 | 1 | 12 |
| Mood-stabilizing | 0 | 1 | 1 | 1 |
| Neuroleptics | 0 | 0 | 0 | 0 |
| Psychostimulants | 0 | 0 | 0 | 0 |

**Note.** Number of patients receiving medication for each therapeutic class in each group (anxiety, no anxiety, and no assessment available). The therapeutic classes are provided for informational purposes only; specific indications are not detailed.

## Notes

### Competing Interest Statement

The authors have declared no competing interest.

### Clinical Trial

NCT01330160

### Author Declarations

The study was conducted between 2011 and 2018 and was approved by the local institutional review board (Comite de Protection des Personnes Nord Ouest IV, Lille, France; reference: 2009-A00141-56, March 17, 2009).

